# Welfare Policies, Joint Pain Prevalence and Educational Gaps in 50 U.S. States from 2011 to 2021: A Fixed Effects Analysis

**DOI:** 10.1101/2025.02.11.25322108

**Authors:** Rui Huang, Yuhang Li, Feinuo Sun

**Affiliations:** University at Buffalo; University at Albany; University of Texas at Arlington

**Keywords:** Welfare Policy, US States, Pain, Education, Disparity

## Abstract

Research on geographic disparities in pain and arthritis-related outcomes is still in its infancy, with little attention to the developing trends over time and the role of state’s welfare policies in shaping pain disparities. This study examines 1) spatiotemporal trends of moderate/severe arthritis-related joint pain prevalence across 50 U.S. states, 2) educational disparities therein, and 3) the impact of welfare policies— i.e., Supplemental Nutrition Assistance Program, Earned Income Credit, minimum wage, unemployment insurance, and Medicaid generosity. This study compiles 6-wave biennial state-level panel data using data from the Behavioral Risk Factor Surveillance System (BRFSS). Logistic regressions are conducted to estimate trends of joint pain prevalence, prevalence for different educational groups (i.e., less than high school, high school or some college, bachelor’s degree or above), and educational disparities. Incorporating policy data from the State Policy & Politics Database (SPPD) and the Kaiser Family Foundation’s database (KFF), fixed effects regressions were used, with state- and year-fixed effects, to assess the impact of welfare policies. Results show that joint pain prevalence has risen in most states, with educational disparities in pain widening in over half, though both trends vary substantially across states. Colorado and North Dakota exhibit considerably sharper increases in both pain prevalence and educational inequalities. Generous Medicaid programs are associated with decreased joint pain prevalence for general population and the least-educated, and marginally linked to smaller educational inequalities therein. This study underscores the importance of state welfare policies in addressing pain disparities and calls for targeted interventions to support less-educated populations.

**Perspectives:** This article draws on state-level panel data and data on state welfare policies to underscore the importance of studying macro-level policy determinants on pain and related disparities.

## 1. Introduction

The prevalence of chronic pain has been rising over the past two decades. ^1^ This trend is concerning, given the strong associations between pain and disability, mortality, and increased healthcare costs. ^2–5^ One major medical condition that causes chronic pain is arthritis, which is highly prevalent in the United States—affecting 23.7% of people (58.5 million) during 2016-2018—with no evidence showing a decrease. ^2,6–10^ Previous studies have documented great educational and geographic disparities in pain and arthritis-related outcomes.^5,11,12^ There is also evidence, though not expansive, showing that state-level policies play a significant role in shaping those outcomes. ^5^ However, most studies on the state-level variations in arthritis-related pain use cross-sectional designs, leaving the developing trends of these outcomes unclear. This paper aims to examine the changes in state-level arthritis-related pain and educational gaps over time, as well as how state policies shape these changes.

Studies on pain from the lens of macro-level determinants are surprisingly limited. According to the eco-social theory of social epidemiology, ^13^ macro-level determinants, including state-level social policies, are crucial in shaping population health. Indeed, extensive empirical studies demonstrate that a state’s higher minimum wage, ^14^ greater Earned Income Tax Credits (EITC), ^15^ Medicaid expansion, ^16^ and more salubrious policies substantially improve population health. Among those studies, only one recent research from Huang et al. ^5^ focused on pain, showing that a more generous Supplemental Nutrition Assistance Program is associated with a lower risk of joint pain. This study, however, relies on cross-sectional data and does not account for temporal changes in both joint pain and state policies. Given that pain and arthritis outcomes develop in different patterns across states and state policies also evolve over the years, leveraging panel data from multiple years can better clarify how the development of arthritis-related pain outcomes is shaped by state policies.

Moreover, the prevalence of pain and arthritis is particularly high among people with low education, who have been shown to be more sensitive to state-level contexts. ^5^ Research shows that the extent of educational disparities in various health outcomes (e.g., sleep, disability, mortality, and arthritis-related pain) vary considerably across states, and the strength of education-health associations is related to state-level welfare provisions. ^5,17–19^ Indeed, by providing more available resources and alleviating psychological strains, welfare policies are crucial for preventing the onset of pain among the most disadvantaged. ^20,21^ However, research focusing on educational disparities in pain barely examines the effects of state welfare policies. Huang et al.’s^5^ study involved some discussions on this topic, but again, it relies on cross-sectional data, and the findings are inconclusive.

This study aims to address gaps in the literature based on a 6-year, biennial, state-level panel dataset covering the years 2011 to 2021, since the Behavioral Risk Factor Surveillance System (BRFSS) provides information about arthritis every other year. The study also include other national data sources, such as the State Policy & Politics Database (SPPD). Focusing on moderate or severe joint pain, this study examines 1) the temporal change of joint pain prevalence from 2011 to 2021 across states, 2) the temporal change of educational disparities in joint pain prevalence in the same period, and 3) how major welfare policies (i.e., SNAP, EITC, minimum wage, unemployment insurance, and Medicaid generosity score) affect arthritis-related joint pain prevalence, and educational disparities therein. Based on findings from previous literature that indicate the associations between welfare policies and health outcomes, ^14–16^ The hypothesize is that more generous *individual* welfare policies are associated with lower prevalence of joint pain, reduced pain prevalence among lower-educated groups, and smaller educational disparities in pain, respectively.

## 2. Data and Methods

This study compiles a panel dataset across 50 U.S. states from multiple data sources (discussed below). The final state-level panel dataset includes 297 state-year observations across six biennial years (2011, 2013, 2015, 2017, 2019, and 2021).

### 2.1 Outcome variables

The outcome variables are constructed from the BRFSS, an annual telephone survey of residents in the U.S. aged 18 and older. It is well-suited for the current research because it gathers comprehensive health-related data and attempts to be representative at the state level. Since 2009, the BRFSS has asked respondents about arthritis and related outcomes every other year. Given that the weighting methods of BRFSS changed in 2011 and questions about arthritis and related symptoms were asked every other year, data were constructed biennially from 2011 to 2021. Thus, six waves of data – data for 2011, 2013, 2015, 2017, 2019, and 2021– were used to construct dependent variables.

The major outcome variable is the prevalence of moderate or severe arthritis-related joint pain (hereinafter “joint pain”) for the total population within each state (denoted as “full”), which is based on questions from BRFSS about doctor-diagnosed arthritis and pain intensity. Respondents were asked to report whether they were diagnosed with “some form of arthritis, rheumatoid arthritis, gout, lupus, or fibromyalgia”. For people who answered “yes” to that question, BRFSS further asked them to rate their joint pain — “During the past 30 days, how bad was your joint pain on average?”—from 0 to 10. Following prior research, ^22^ a dichotomized measure is generated, with a score of 6 or higher indicating “moderate or severe joint pain.” This study focuses on moderate or severe pain because this level of pain intensity is more clinically relevant and has been proven to be strongly linked to functional limitation and mortality. ^23,24^

In addition, four outcomes are constructed to examine educational disparities in arthritis-related joint pain: the prevalence of joint pain among individuals with less than a high school degree (“<HS”), those with a high school or some college education (“HS/SC”), those with a bachelor’s degree or higher (“BA+”), and the educational gap between <HS and BA+ groups within each state (“educational gap”). The use of a three-category measure of educational attainment aligns with prior research. ^5,25^

The analytical sample is restricted to respondents aged 25 years old or above since most people complete their education at this age. ^17^ Observations without corresponding state-year information are also deleted. BRFSS Data from New Jersey in 2019 and Florida in 2021 were excluded because they did not meet the minimum requirement of BRFSS for inclusion. ^26,27^ This results in a 2,531,412 sample size. The outcome variable, moderate/severe joint pain, was missing in 2.75% of cases, and other covariates (i.e., age, sex/gender, race/ethnicity, education) were missing in 0.01% to 1.45%. 5.88% of participants in total have missing data on variables of interest. 2,382,564 cases are finally obtained after deleting these observations. The sample size for each state each year is shown in the Supplementary Table S1

The pain prevalence was firstly estimated using logistic regression models for each year where state dummies interact with age, sex/gender and race/ethnicity, as this study assumes that the effects of these factors vary by state contexts. Then, the prevalence for three educational groups were estimated using similar models, but respondent’s education was also included as a predictor. All models were adjusted by final sample weights, and the prevalence was calculated using margins command in Stata. In order to examine the policy effects on the educational gap, an “educational gap” variable (i.e., educational gap = the prevalence for <HS – the prevalence for BA+) were constructed. Data for Alaska in 2015 were excluded since the prevalences show evident inconsistency (i.e., the prevalences are all less than 1%). As a result, 297 state-year data points from the BRFSS were available for analysis.

### 2.2 Key independent variables

The impacts of five welfare policies were examined: Supplemental Nutrition Assistance Program (SNAP), Earned Income Tax Credit (EITC), minimum wage, Unemployment insurance (UI), and Medicaid Generosity Score (MGS). These policies differ in terms of policy type, benefit form, and target population. ^28^ They constitute the major social safety net for people with limited access to resources across various domains, including health behavior (influenced by SNAP), tax (via EITC), labor law and benefits (through minimum wage and UI benefits), and health care (through Medicaid generosity). All of them have been shown to improve health outcomes for socioeconomic-disadvantaged people significantly. ^28–31^ Data for SNAP are from the Kaiser Family Foundation’s database (KFF) and other four state-level predictors (i.e., EITC, minimum wage, UI, and MGS) from the State Policy & Politics Database (SPPD). Measures of these policies are as follows:

1. SNAP is measured by average monthly benefits (adjusted in 2021 dollars) received per participant.
2. EITC is measured by state EITC rate as the percentage of federal credit.
3. The minimum wage is measured by state minimum wage, adjusted in 2021 dollars.
4. Unemployment insurance is measured by the maximum unemployment insurance benefit that eligible individuals can receive, adjusted in 2021 dollars.
5. Medicaid generosity score is an index. This comprehensive measure is constructed by Fox and colleagues. ^32^ and capture four major dimensions of Medicaid program, including (1) income eligibility (i.e., income eligibility thresholds for different age categories of children, pregnant women, parents, and nonparents), (2) immigrant benefits (i.e., whether the state provides Medicaid to lawful and/or unauthorized immigrants during their first five years), (3) administrative burden (e.g., asset tests, whether applying by phone, waiting period, etc.), and (4) benefit levels (e.g., whether Medicaid covers dentist, eye glasses, psychologist, etc.). All underlying variables were assigned equal weight, summed, and normalized to a 0–100 scale, where 0 represents the least generous and 100 the most generous. Details on how each underlying variable was evaluated and scored can be found in Fox et al.’s article.^33^

Following prior studies,^31^ these policy factors are drawn from the two years prior to the outcome variables (2009, 2011, 2013, 2015, 2017, and 2019) to account for the time lag in social policy impacts. There are other studies using different periods depending on their research contexts.^28,34–36^ In the current analysis, while models with two-year lags provided the most consistent and interpretable findings and the findings are somewhat consistent across models with different lag structures (discussed in the Result section below), we acknowledge that this lag structure was selected empirically. The full set of alternative lag models is presented in the Supplemental Table S5 for transparency. A correlation matrix for policy variables is summarized in Supplementary Table S2.

### 2.3 Covariates

Several state-level covariates were also considered, including urbanization rate, the percentage of immigrants, gross domestic product (GDP) growth rate, and income inequality (measured by the Gini index). Research has shown that the prevalence of chronic pain linearly increases from metropolitan to nonmetropolitan areas in the US.^37^ At the same time, rural areas tend to have higher proportion of Republican voters, who often hold less favorable view towards expanding governmental aid.^38^ Additionally, GDP growth rate, as a comprehensive indicator of changes in macro-economic environment, may have significant impacts on both the generosity of welfare policies in a state and the risk of chronic pain among its residents, potentially through pathways such as stress. Moreover, income inequality has been shown to be associated with both welfare generosity and educational disparity in pain.^5,39^ The analysis also controlled for the percentage of immigrants, given that immigrants tend to have a better health^40^ and states with more immigrants may have lower pain prevalence. Data on the percentage of immigrants are drawn from the mid-year American Community Survey (ACS) 5-year estimates. GDP growth data are from the U.S. Bureau of Economic Analysis. Descriptive statistics for all variables are shown in Supplementary Table S3.

### 2.4 Analytical Strategies

The unit of analysis is “state-year”. One figure was plotted showing the trends of joint pain prevalence over years as well as geographic distributions of the relative change in joint pain prevalence and educational gaps between 2011 and 2021, defined as *(pain2021 – pain_2011_)/pain_2011_*, across states. Second, two-way fixed effects regression models are conducted to examine the association between state policy factors and five outcomes, including joint pain prevalence for the full population and for three educational groups, as well as the educational gap in joint pain prevalence, controlling for state-level covariates (Models 1-5). To account for temporal dependencies, year-fixed effects were incorporated in all models. This controls for unmeasured secular trends that affect the entire nation (e.g., changes in national economy and policy changes at the federal level). Additionally, state-fixed effects were included to account for time-invariant unmeasured factors across states (e.g., population composition, stable difference in policy by state, cost of living, climate and environment features, etc.).^41,42^ Spatial panel fixed-effect models were also explored to account for both spatial and temporal dependencies (results are available upon request) but the spatial parameter was not significant and the results are not interpretable, suggesting that these models do not fit the data in current study. All variables are log-transformed except for the GDP growth rate, which contains negative values. Because the EITC variable includes zero values for some cases, we added a small constant – i.e., log(EITC +1) – when EITC = 0 to enable log transformation. Thus, estimates represent the corresponding percentage change in each type of pain measure for each 1% increase in each type of welfare policy. The analysis also clustered the standard errors at the state level to screen out the potential problems that may be caused by heteroscedasticity or serial correlation. Variance inflation factors (VIFs) show that multicollinearity is not a problem. Equations for Models 1-5 are shown in the Supplementary. All models were analyzed in Stata 17.0.

Additional analyses were conducted to test the sensitivity of results. Instead of pooling all policy indicators together, several sets of models were estimated: (1) models where policy variables are included one at a time without state-level covariates, (2) similar models that incorporate state-level covariates, and (3) models where the full set of policy indicators and state-level covariates are included, and pain outcomes are lagged by zero, 1, 3, 4 years. These results are presented in Supplementary Tables S4 and S5.

All Stata code used to clean the data, construct variables, and estimate models is available upon request. Data are accessible through the original provider listed above. All data used in this study are publicly available and deidentified, and thus this research does not need to be reviewed by the Institutional Review Board.

## 3. Results

### 3.1 Temporal Trends of Joint Pain Prevalences and Educational Disparities

Figure 1 shows the trends of joint pain prevalence for the full population and for three educational groups, as well as educational gaps from 2011 to 2021. Results indicate a steady rise in the joint paint prevalence (from 10.5% to 11.6%; the solid red line) across the entire U.S. population. Similarly, the prevalences for three different educational groups (i.e., <HS, HS/SC, BA+) tend to increase over time. The trend in the educational gap (the solid orange line) shows fluctuations, with a notable spike in 2017.

**Figure 1.**
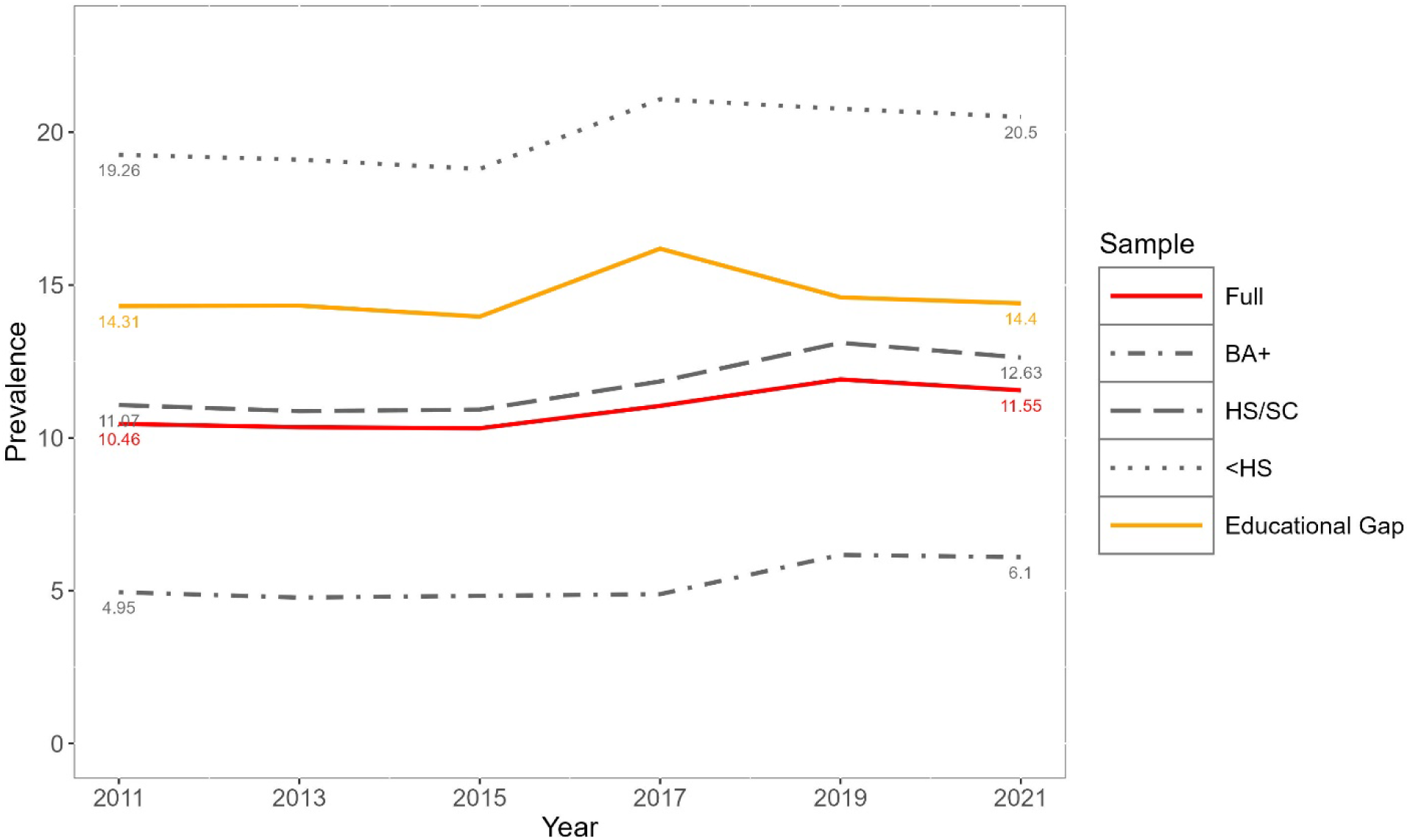
Trend of moderate/severe joint pain prevalence, prevalence for educational groups, and educational gap between less than high school and bachelor’s degree or above. Note: a. Prevalences are adjusted by age, sex/gender, race/ethnicity. b. Full = prevalence for full sample (without differentiating education); BA+ = prevalence for bachelor’s degree or above; HS/SC = prevalence for high school or some college; &lt;HS = prevalence for less than high school; Educational Gap = prevalence for less than high school – joint pain prevalence for bachelor’s degree or above.

Figures 2 and 3, respectively, demonstrate the geographic distributions of relative change (i.e., percent change rates) in overall joint pain prevalence and educational gaps across states between 2011 and 2021. Corresponding trend figures are presented in Supplementary Figures S1 and S2. Figure 2 illustrates that joint pain prevalence increases in most states and the average increasing rate is 11.0% (SD = 7.5%). However, the increasing rates vary considerably across states. The rate of increase was highest in Colorado (26.2%), followed by Hawaii, Nebraska, Kansas, and North Dakota. Only four states – Montana, Wyoming, South Dakota, and New York – exhibit a mild decreasing trend (less than 2.2%).

**Figure 2.**
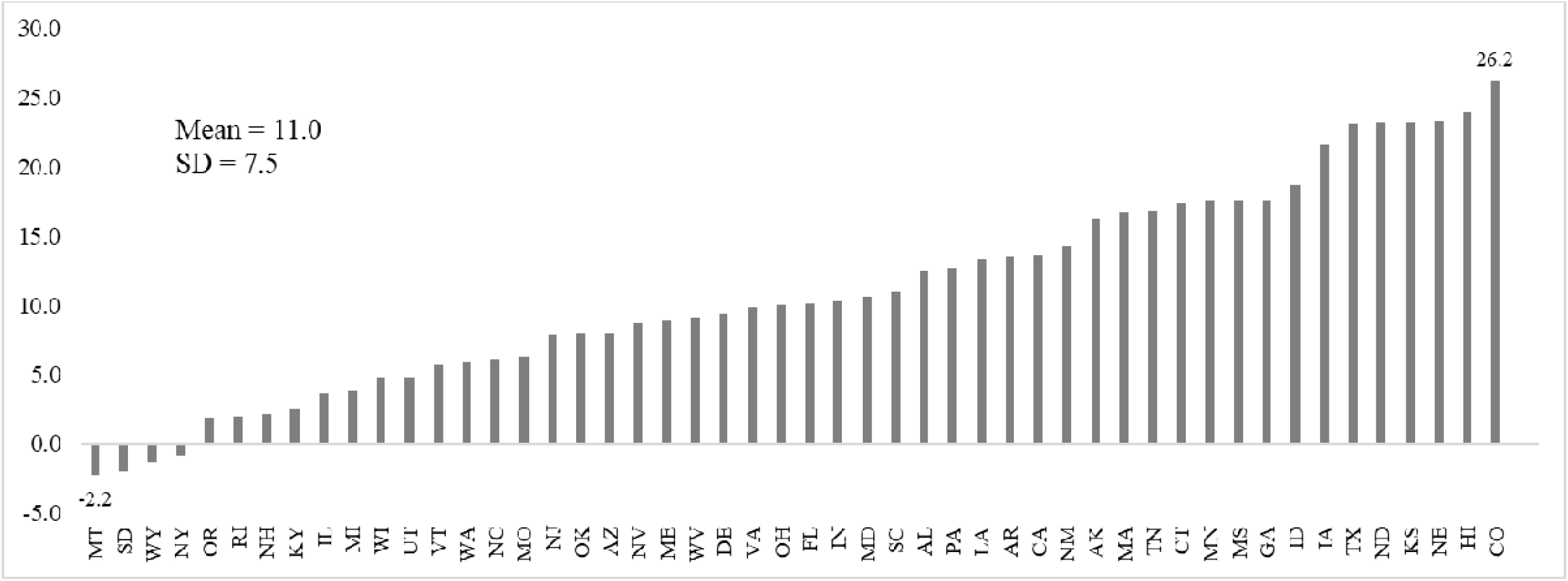
Relative prevalence changes among U.S. adults between 2011 and 2021, by state. Note: a. The prevalence change for Florida is from data between 2011 and 2019 since data from Florida did not meet the minimum requirements of BRFSS for inclusion. b. Percentage change rate = (Pain_2021_ – Pain_2011_)/Pain_2011_.

**Figure 3.**
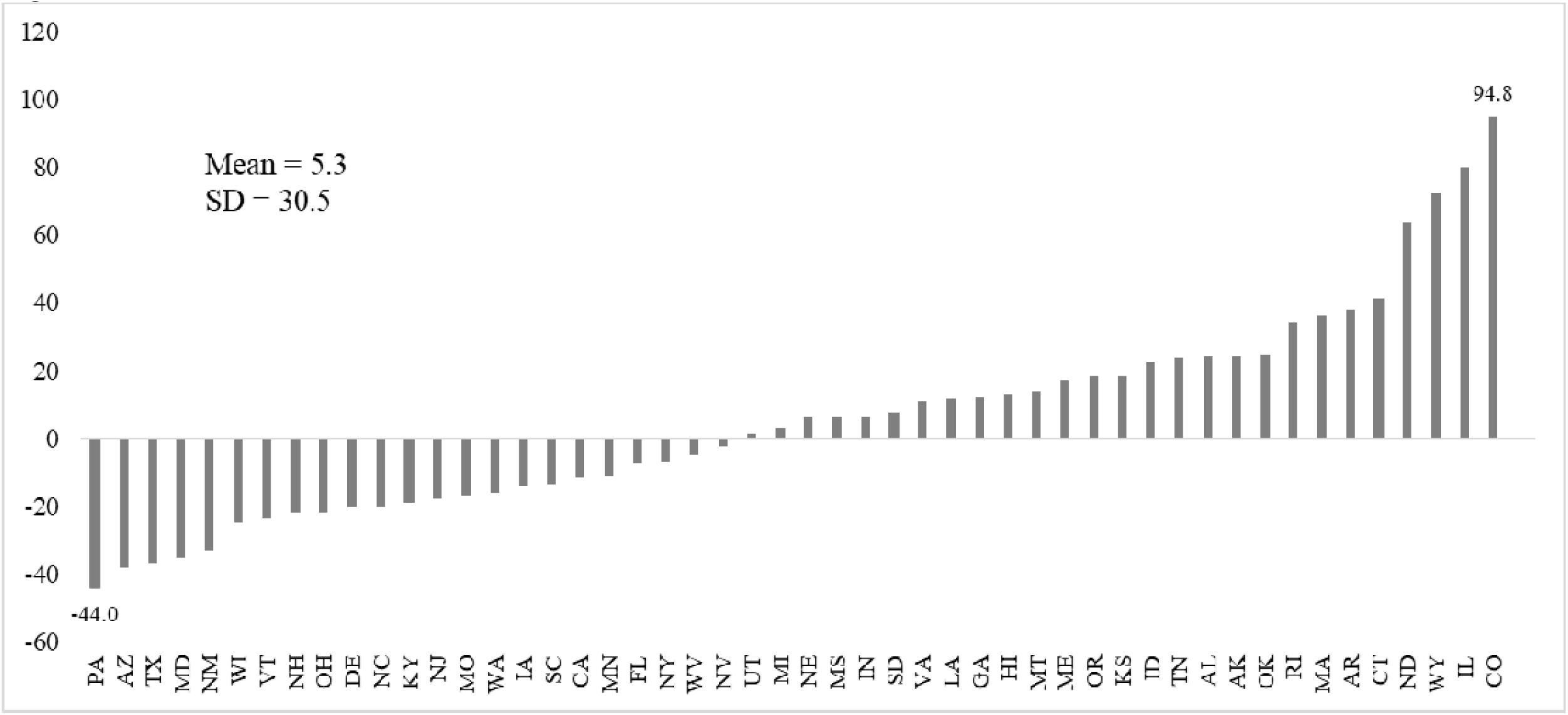
Relative prevalence changes in educational gaps among U.S. adults between 2011 and 2021, by state. Note: a. The prevalence change for Florida is from data between 2011 and 2019 since data from Florida did not meet the minimum requirements of BRFSS for inclusion. b. Percentage change rate = (Educational Gap_2021_ – Educational Gap_2011_)/Educational Gap_2011_.

Figure 3 shows that the average increasing rate of educational gap in joint pain is 5.3%, with substantial variation across states (SD = 30.5). More than half of the 50 states experienced a rise in the educational gap, while the other half saw a decline. The highest increase in the educational gap was observed in Colorado (94.8%), followed by Illinois, Wyoming, North Dakota, and Connecticut. In contrast, Pennsylvania experienced the most significant decrease (44.0%), followed by Arizona, Texas, Maryland, and New Mexico.

### 3.2 The Effect of Welfare Policies

Table 1 summarizes the results from two-way fixed effect models and the effects of welfare policies on different measures of joint pain prevalence, controlling state-level covariates. Given that standard errors are clustered and the sample size is relatively small (N=297), results are presented at both at the 95% confidence level and at the more relaxed 90% level. The goal of this analysis is to identify potentially meaningful associations and avoid the risk of underestimating the impacts. Findings at the 90% level are interpreted cautiously as marginally significant and are explicitly labeled as such in the text. This study emphasizes effect sizes and/or associations rather than statistical significance alone.

**Table 1.**
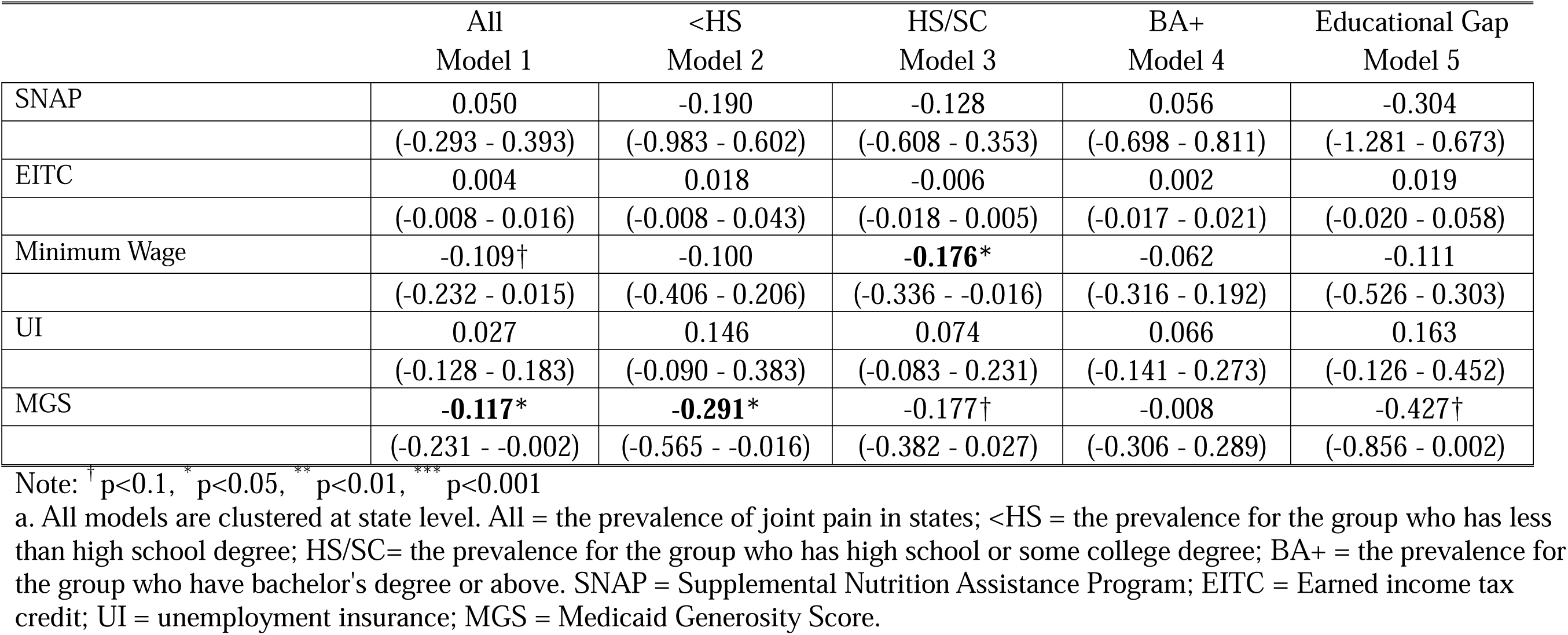
Coefficients of welfare policies in two-way fixed effects models.

|  | All<br>Model 1 | <HS<br>Model 2 | HS/SC<br>Model 3 | BA+<br>Model 4 | Educational Gap<br>Model 5 |
| --- | --- | --- | --- | --- | --- |
| SNAP | 0.050<br>(-0.293 - 0.393) | -0.190<br>(-0.983 - 0.602) | -0.128<br>(-0.608 - 0.353) | 0.056<br>(-0.698 - 0.811) | -0.304<br>(-1.281 - 0.673) |
| EITC | 0.004<br>(-0.008 - 0.016) | 0.018<br>(-0.008 - 0.043) | -0.006<br>(-0.018 - 0.005) | 0.002<br>(-0.017 - 0.021) | 0.019<br>(-0.020 - 0.058) |
| Minimum Wage | -0.109†<br>(-0.232 - 0.015) | -0.100<br>(-0.406 - 0.206) | **-0.176\***<br>(-0.336 - -0.016) | -0.062<br>(-0.316 - 0.192) | -0.111<br>(-0.526 - 0.303) |
| UI | 0.027<br>(-0.128 - 0.183) | 0.146<br>(-0.090 - 0.383) | 0.074<br>(-0.083 - 0.231) | 0.066<br>(-0.141 - 0.273) | 0.163<br>(-0.126 - 0.452) |
| MGS | **-0.117\***<br>(-0.231 - -0.002) | **-0.291\***<br>(-0.565 - -0.016) | -0.177†<br>(-0.382 - 0.027) | -0.008<br>(-0.306 - 0.289) | -0.427†<br>(-0.856 - 0.002) |

Note: † p<0.1, \* p<0.05, \*\* p<0.01, \*\*\* p<0.001
a. All models are clustered at state level. All = the prevalence of joint pain in states; <HS = the prevalence for the group who has less than high school degree; HS/SC= the prevalence for the group who has high school or some college degree; BA+ = the prevalence for the group who have bachelor's degree or above. SNAP = Supplemental Nutrition Assistance Program; EITC = Earned income tax credit; UI = unemployment insurance; MGS = Medicaid Generosity Score.

Results demonstrate that net of other factors, a 1% increase in Minimum wage is associated with 0.18% (β: -0.176, 95% Confidence Interval [CI]: -0.336 – -0.016, p = 0.032) a decrease in joint pain prevalence for the HS/SC group. A 1% increase in Medicaid Generosity Score (MGS) is related to a 0.12% decrease in overall joint pain prevalence (β: -0.117, 95%CI: - 0.231 - -0.002, p = 0.047). This increase also corresponds to a 0.29% reduction in joint pain prevalence of <HS group (β:-0.291, 95%CI: -0.565 – -0.016, p =0.038), a 0.18% reduction in the prevalence of HS/SC group (β: -0.177, 95% CI: -0.382 – 0.027, p = 0.088), and a narrowing of educational disparity therein (β: -0.427, 95%CI: -0.856 – 0.002, p = 0.051), though the statistical significance for the HS/SC group and educational disparity is marginally significant and should be interpreted with caution.

Findings from additional analyses show that models examining policy indicators separately (Table S4) yield results similar to the saturated models in Table 1. Results from models with different lag periods (Table S5) suggest that a two-year lag in pain outcomes is a more appropriate and conservative approach, as welfare policies exhibit little impact with 0- or 1-year lags but show increasing effects in models with 3- or 4-year lags.

## 4. Discussion

Arthritis-related joint pain is a costly and prevalent condition that seriously compromises the quality of life in the U.S. .^2,43^ Data of this study show that the prevalence of moderate to severe joint pain has increased over the past decade, rising from 10.45% in 2011 to 11.54% in 2021 – around 4.6 million more people were affected. Joint pain prevalence across educational groups (<HS, HS/SC, BA+) has increased, each following distinct temporal patterns. The trend of educational gap fluctuated over time, with a notable spike in 2017, which is primarily driven by a substantial increase in joint pain prevalence among those with less than a high school education. In contrast, the decline in the gap observed in 2019 is largely attributable to a rising prevalence among individuals with a bachelor’s degree or higher.

Although macro-level factors and cross-state disparities are critical for understanding the development of health inequalities, ^18,44^ to our knowledge, this is the first study documenting the temporal trend of pain and educational disparities therein across states and examining policy determinants on the developing trends of pain over time.

Results reveal that the developing trends of joint pain prevalence from 2011 to 2021 vary substantially across 50 states. The prevalence increases in most states except for Montana, Wyoming, South Dakota, and New York. The increasing trends in Colorado, Hawaii, Nebraska, Kansas, and North Dakota are sharper (Figure 2). Moreover, there are substantial state-level variations in changes of educational disparities, with the educational gap increasing in half of the states and decreasing in the other half. Colorado had the highest increase in the educational gap (94.8%), followed by Illinois, Wyoming, North Dakota, and Connecticut, while Pennsylvania saw the largest decrease (44.0%), followed by Arizona, Texas, Maryland, and New Mexico. Both Colorado and North Dakota exhibit sharper increases in both joint pain prevalence and educational disparities due to the notably high increase in joint pain prevalence among the <HS group (relative increases are 63.1% in Colorado and 47.0% in North Dakota, respectively; not shown but available by request). This aligns with previous studies showing that substantial state-level differences in health outcomes are primarily driven by disparities among the least-educated populations. ^5,44^ Policy interventions are urgently needed in these two states to halt the disproportionate increase in pain prevalence.

Another major goal of this study is to examine the impacts of welfare policies. Results reveal that the increase in minimum wage is associated with a decreased prevalence for the HS/SC group. However, this positive impact does not appear in the least-educated group (<HS) nor the educational gap in pain. The impact of minimum wage on health is inconclusive: while some studies suggest that a higher minimum wage improves health outcomes, ^45–47^ others report no effects or mixed and conflicting results. ^48–51^ Some evidence also points to a positive association between a higher minimum wage and unhealthy behaviors such as smoking, poor diets and obesity ^52,53^, though the opposite findings exist as well. Findings of this study highlight the critical role of education in this particular dynamic: better education may enable individuals to make better (and healthier) use of additional disposable income.

In addition, findings of current study reveal that the increase in the MGS is linked to favorable results in terms of reduced overall pain prevalence and lower prevalence for the least-educated group (i.e., < HS), and is marginally associated with lower prevalence for the HS/SC group and a narrower educational gap. Unlike other policies that show limited effects on eliminating inequalities in this study and other places, ^54^ Medicaid, a healthcare safety net program, shows effective influences on tackling health inequities through both “resources” and “psychology” domains proposed by the Fundamental Cause Theory. ^55,56^ In addition to increasing access to higher-quality care,^57^ a more generous Medicaid program serves as a vital resource in alleviating the financial burden and psychological distress faced by the less-educated.^29,58,59^ All of them can collectively empower residents to better manage arthritis and control pain.

While the estimated effects may appear small in absolute percentage terms (e.g., a 0.12% decrease in joint pain prevalence per 1% increase in MGS), they have important implications at the population level. Given that joint pain affects 23.7% of U.S. adults (58.5 million),^43^ even a 0.12% relative reduction can translate to around 70,000 fewer individuals experiencing pain (corresponding to β = -0.117 in Table 1) when applied to the adult population if Medicaid generosity increases slightly – i.e., 1%. Similarly, a 1% increase in minimum wage could potentially benefit around 68,000 individuals with HS/SC degree (corresponding to β = -0.176 in Table 1; equations are available upon request). Since pain is leading cause of disability and a major driver of healthcare expenditure, even small proportional decreases can substantially improve more people’s well-being reduce the broader societal burdens in terms of healthcare utilization and productivity losses. ^4,60^ It is important to note that, to avoid ecological fallacy, state-level effects cannot be directly interpreted as individual-level risk reductions.

Significant effects of SNAP were not observed, EITC, and UI, suggesting that some seemingly salubrious policies may fail to reduce pain risks. Supplemental analyses indicate that these nonsignificant findings are affected little by the inclusion of state-level covariates or the time lags used (see Tables S4 and S5). There are multifaceted reasons. For instance, welfare participation is often associated with stigma that adds extra psychological burdens to recipients.^61^ Research shows that while SNAP can promote self-rated health and more utilization of healthcare resources, it is also suggested to be related to obesity, another strong predictor of chronic pain.^62,63^ Therefore, some policies could generate benefits while opening up new pathways toward higher risks of pain. Note that due to the relatively small number of panel years, the findings of this study might be subject to Type II error. Studies investing effects of policy factors on pain are extremely limited and there is only one previous study from Huang et al.^5^ examining the effects of SNAP and EITC on joint pain. Consistent with the current study, they did not find a significant relationship between EITC and pain. While a link between SNAP and lower risk of pain has been reported, SNAP cannot alleviate educational disparities therein, which is also consistent with findings of this study. However, since their study relied on cross-sectional models, the findings are more susceptible to confounding factors compared to the current analysis, which employs multi-wave data and fixed effects models. It is also important to bear in mind that the lack of association between these measures and pain-related outcomes should not be overinterpreted; it only implies these measures of policies are unimportant in shaping the change of state-level pain prevalences. More research is needed with available data.

This study is subject to several limitations. First, the use of state-level data only allows us to conclude the effects of policies on the population level. Making implications on other levels, such as individual-level pain risk, would commit to ecological fallacy. More research is needed on different levels of analysis. Second, although several covariates were included, findings this study may still suffer from bias resulting from omitted time-varying variables, such as cultures and government ideology, which calls for further analysis when more data are available. Third, social policies often come in a package and interact with each other in complicated ways that are often dependent on other contextual factors. ^28,64^ Future research needs to disentangle the interactions between different state policies and social-environmental factors. Fourth, some of the interpretations include estimates using a 90% confidence level, which may increase the risk of Type I error. However, rather than confirmatory testing, the goal of this analysis is to identify potentially meaningful associations and avoid underestimating important effects. This is consistent with previous policy evaluation studies.^31,65^

In conclusion, this study underscores the importance of state-level macro factors by showing large heterogeneity of temporal trends of pain prevalence and disparities across U.S. states during the past decades. Colorado and North Dakota are particularly disadvantageous with fast-rising joint pain prevalence and educational inequality therein. Urgent interventions are needed as the less-educated people in these two states appear to be more vulnerable and are likely to experience more pain-related health issues (e.g., disability) in the future. Moreover, higher minimum wages and more generous Medicaid are effective in reducing the population’s pain and educational disparities. This study highlights that future studies on pain should move beyond the individual level and identify macro-level barriers to effective pain prevention or management. Investigating welfare policies offers an important perspective in exploring how macro sociopolitical contexts shape the mechanisms contributing to pain inequality.

## Data Availability

All data produced in the present study are available upon reasonable request to the authors.

https://www.cdc.gov/brfss/data_documentation/index.htm

https://asi.syr.edu/state-policy-politics-database-sppd/

https://www.kff.org/other/state-indicator/total-snap-program-benefits/?currentTimeframe=0&sortModel=%7B%22colId%22:%22Location%22,%22sort%22:%22asc%22%7D

https://www.census.gov/programs-surveys/acs

https://www.bea.gov/data/gdp/gdp-state

## Acknowledgements

The authors would like to thank Anna Zajacova for her comments on the manuscript.

## Author contributions

**Rui Huang**: Conceptualization, Methodology, Validation, Formal analysis, Investigation, Resources, Data Curation, Writing - Original Draft, Writing - Review & Editing, Visualization, Project administration. **Yuhang Li**: Methodology, Validation, Resources, Writing - Review & Editing, Visualization. **Feinuo Sun**: Conceptualization, Methodology, Validation, Writing - Review & Editing, Supervision, Funding acquisition.

## Data availability

Data and Stata code are available by request.

