## Supplementary for "Welfare Policies, Joint Pain Prevalence and Educational Gaps in 50 U.S. States from 2011 to 2021: A Fixed Effects Analysis"

**Supplementary material**

1. Two-way fixed effects models estimating the associations between individual welfare policies and arthritis-related joint pain prevalence by the overall population, different educational groups, and educational gap (Models 1-5).

$${lnY}_{st}= \beta_{0}+ \beta_{1}{lnSNAP}_{s,t-2}+ \beta_{2}{lnEITC}_{s,t-2}+ \beta_{3}{\ln(Minimum Wage)}_{s, t-2}+ \beta_{4}{lnUI}_{s,t-2}+ \beta_{5}{lnMGS}_{s, t-2}+ \beta_{k}{lnCovariates}_{st}+ \alpha_{s}+ \gamma_{t}+ \varepsilon_{st}$$

Where s represents state, t = 2011, 2013…2019, 2021, represents year. $Y_{st}$ denotes the log-transformed arthritis-related joint pain prevalence (varies by model) in state *s* and year t; k = 1…k, represents number of covariates; $\alpha_{s}$ represents state-fixed effects; $\gamma_{t}$ represents year-fixed effects; $\varepsilon_{st}$ represents error term (clustered at the state level).

Model 1: $Y_{st}=$overall joint pain prevalence. Model 2: $Y_{st}=$ joint pain prevalence for the group with less-than-high-school degree.

Model 3: $Y_{st}=$ joint pain prevalence for the group with high school or some college.

Model 4: $Y_{st}=$ joint pain prevalence for the group with bachelor’s degree or above.

Model 5: $Y_{st}=$ educational gap between <HS and BA+.

Table S1. Sample sizes for each state each year for estimating the prevalence of arthritis-related joint pain.

| State name | 2011 | 2013 | 2015 | 2017 | 2019 | 2021 |
| --- | --- | --- | --- | --- | --- | --- |
| Alabama | 6,927 | 5,886 | 6,918 | 6,141 | 6,466 | 4,185 |
| Alaska | 3,119 | 3,982 | . | 2,876 | 2,655 | 4,806 |
| Arizona | 5,896 | 3,714 | 6,963 | 13,818 | 8,018 | 9,510 |
| Arkansas | 4,271 | 4,703 | 4,656 | 4,947 | 4,913 | 4,875 |
| California | 15,344 | 9,958 | 10,733 | 8,205 | 10,021 | 5,877 |
| Colorado | 12,316 | 11,997 | 11,619 | 8,645 | 8,187 | 9,105 |
| Connecticut | 6,224 | 6,845 | 10,459 | 9,578 | 8,240 | 7,355 |
| Delaware | 4,370 | 4,659 | 3,543 | 3,670 | 3,479 | 3,251 |
| Florida | 11,013 | 29,177 | 8,363 | 19,623 | 15,123 | . |
| Georgia | 9,069 | 7,049 | 4,150 | 5,344 | 6,495 | 7,271 |
| Hawaii | 6,954 | 6,903 | 6,368 | 7,026 | 6,982 | 7,008 |
| Idaho | 5,477 | 4,777 | 5,054 | 4,406 | 4,810 | 5,916 |
| Illinois | 5,051 | 5,078 | 4,735 | 5,109 | 4,947 | 2,739 |
| Indiana | 7,510 | 8,352 | 5,283 | 12,564 | 7,947 | 8,761 |
| Iowa | 6,586 | 7,379 | 5,553 | 6,907 | 8,758 | 8,361 |
| Kansas | 19,136 | 20,955 | 20,351 | 19,458 | 10,074 | 15,705 |
| Kentucky | 9,618 | 9,771 | 7,627 | 7,758 | 7,188 | 4,797 |
| Louisiana | 10,016 | 4,782 | 4,118 | 4,300 | 4,190 | 4,540 |
| Maine | 12,180 | 6,938 | 8,105 | 9,051 | 10,334 | 10,633 |
| Maryland | 9,239 | 11,786 | 11,175 | 12,392 | 15,801 | 13,885 |
| Massachusetts | 19,871 | 13,173 | 7,782 | 6,015 | 6,611 | 6,486 |
| Michigan | 9,970 | 11,198 | 7,793 | 9,634 | 9,329 | 8,338 |
| Minnesota | 13,896 | 12,944 | 14,885 | 15,335 | 13,797 | 14,189 |
| Mississippi | 8,063 | 6,687 | 5,490 | 4,629 | 4,684 | 3,954 |
| Missouri | 5,808 | 5,704 | 6,427 | 6,794 | 6,549 | 10,769 |
| Montana | 9,344 | 8,727 | 5,448 | 5,362 | 5,847 | 5,548 |
| Nebraska | 23,054 | 15,519 | 15,732 | 14,037 | 14,534 | 13,454 |
| Nevada | 4,839 | 4,519 | 2,573 | 3,386 | 2,422 | 2,387 |
| New Hampshire | 5,814 | 5,806 | 6,304 | 5,315 | 5,383 | 5,976 |
| New Jersey | 13,813 | 11,677 | 10,054 | 10,642 | . | 6,797 |
| New Mexico | 8,508 | 8,130 | 5,960 | 5,920 | 5,378 | 5,614 |
| New York | 6,678 | 7,593 | 10,157 | 10,688 | 12,438 | 34,367 |
| North Carolina | 10,535 | 7,958 | 5,881 | 4,382 | 3,762 | 4,389 |
| North Dakota | 4,781 | 6,522 | 4,298 | 6,402 | 5,088 | 5,292 |
| Ohio | 8,999 | 10,668 | 10,683 | 11,261 | 12,302 | 12,803 |
| Oklahoma | 7,888 | 7,517 | 6,280 | 6,013 | 5,645 | 4,773 |
| Oregon | 5,579 | 5,209 | 4,623 | 4,714 | 5,294 | 4,617 |
| Pennsylvania | 10,138 | 9,325 | 4,978 | 5,803 | 5,867 | 5,502 |
| Rhode Island | 5,939 | 5,731 | 5,411 | 5,131 | 5,567 | 4,997 |
| South Carolina | 11,467 | 9,477 | 10,203 | 10,213 | 6,335 | 9,108 |
| South Dakota | 7,625 | 6,201 | 6,605 | 6,488 | 6,004 | 6,557 |
| Tennessee | 5,423 | 5,115 | 5,243 | 5,237 | 5,582 | 4,279 |
| Texas | 13,488 | 9,455 | 12,606 | 10,987 | 10,887 | 9,433 |
| Utah | 11,249 | 11,188 | 9,916 | 8,846 | 10,256 | 9,237 |
| Vermont | 6,528 | 5,767 | 5,772 | 5,921 | 5,923 | 6,022 |
| Virginia | 5,931 | 7,444 | 7,697 | 8,686 | 8,848 | 8,848 |
| Washington | 13,618 | 10,001 | 14,153 | 11,844 | 11,653 | 11,813 |
| West Virginia | 4,883 | 5,374 | 5,264 | 5,090 | 4,847 | 6,150 |
| Wisconsin | 4,728 | 5,728 | 5,427 | 5,272 | 4,523 | 5,573 |
| Wyoming | 6,175 | 5,830 | 4,933 | 4,098 | 4,208 | 3,999 |
| Total | 444,948 | 420,878 | 376,733 | 395,963 | 364,191 | 379,851 |

Note: BRFSS Data from New Jersey in 2019 and Florida in 2021 are excluded because they did not meet the minimum requirement of BRFSS for inclusion. Data for Alaska in 2015 are excluded since the prevalence estimates are implausibly low (i.e., the prevalences are all less than 1%).

Table S2. Correlation matrix of state-level policy variables.

|  | SNAP | EITC | Minimum wage | UI |
| --- | --- | --- | --- | --- |
| EITC | -0.00 |  |  |  |
| Minimum wage | 0.23 | 0.37 |  |  |
| UI | 0.10 | 0.26 | 0.40 |  |
| MGS | 0.15 | 0.50 | 0.50 | 0.53 |

Note: SNAP = Supplemental Nutrition Assistance Program; EITC = Earned Income Tax Credit; UI = unemployment insurance; MGS = Medicaid Generosity Score.

Table S3.  Descriptive statistics (N=297)

| *Arthritis-related Joint pain prevalences (%)* | Mean | SD |
| --- | --- | --- |
| All | 10.90 | (2.85) |
| <HS | 19.91 | (5.37) |
| HS/SC | 11.73 | (2.59) |
| BA+ | 5.28 | (1.42) |
| Educational Gap | 14.64 | (4.76) |
| *Welfare policies* |  |  |
| SNAP ($) | 147.91 | (21.55) |
| EITC (%) | 8.08 | (12.26) |
| Minimum wage ($) | 9.08 | (1.12) |
| Unemployment insurance ($) | 13,589.55 | (4,917.05) |
| Medicaid Generosity Score | 52.87 | (10.08) |
| *State-level covariates* |  |  |
| Urbanization rate (%) | 72.90 | (14.54) |
| Gini | 0.46 | (0.02) |
| GDP growth rate | 2.17 | (2.20) |
| Percentage of immigrants (%) | 9.09 | (6.02) |

a. Joint pain prevalences are adjusted for age, sex, race/ethnicity. All estimates are sample weight adjusted.

b. All = states’ prevalence of joint pain; <HS = the prevalence for the group who has less than high school degree; HS/SC= the prevalence for the group who has high school or some college degree; BA+ = the prevalence for the group who have bachelor's degree or above. Educational gap = the prevalence for <HS – the prevalence for BA+.

c. Data on arthritis-related joint pain prevalence and state-level covariates are from years 2011, 2013, 2015, 2017, 2019, and 2021; data on welfare-policies are from years 2009, 2011, 2013, 2015, 2017, and 2019

Table S4. Coefficients of welfare policies in two-way fixed effects models for each policy with two-year lags.

|  | All | <HS | HS/SC | BA+ | Educational Gap |
| --- | --- | --- | --- | --- | --- |
| Panel A |  |  |  |  |  |
| SNAP | 0.042 | -0.325 | -0.214 | -0.075 | -0.437 |
|  | (-0.309 - 0.394) | (-1.118 - 0.469) | (-0.701 - 0.272) | (-0.903 - 0.754) | (-1.405 - 0.532) |
| EITC | 0.002 | 0.019 | -0.008 | 0.002 | 0.019 |
|  | (-0.010 - 0.013) | (-0.009 - 0.046) | (-0.020 - 0.004) | (-0.016 - 0.020) | (-0.024 - 0.062) |
| Minimum Wage | -0.086 | -0.029 | -0.151† | -0.027 | -0.038 |
|  | (-0.207 - 0.035) | (-0.323 - 0.265) | (-0.308 - 0.005) | (-0.263 - 0.209) | (-0.439 - 0.363) |
| UI | 0.043 | 0.200† | 0.06 | 0.065 | 0.240† |
|  | (-0.093 - 0.178) | (-0.012 - 0.412) | (-0.074 - 0.193) | (-0.110 - 0.239) | (-0.028 - 0.508) |
| MGS | -0.078 | -0.254† | -0.182† | -0.059 | -0.348† |
|  | (-0.173 - 0.016) | (-0.517 - 0.009) | (-0.386 - 0.023) | (-0.364 - 0.245) | (-0.735 - 0.039) |
| Panel B |  |  |  |  |  |
| SNAP | 0.041 | -0.255 | -0.134 | 0.035 | -0.375 |
|  | (-0.326 - 0.408) | (-1.116 - 0.606) | (-0.665 - 0.397) | (-0.748 - 0.819) | (-1.430 - 0.680) |
| EITC | 0.002 | 0.021 | -0.007 | 0.004 | 0.022 |
|  | (-0.010 - 0.014) | (-0.009 - 0.050) | (-0.019 - 0.005) | (-0.013 - 0.020) | (-0.021 - 0.065) |
| Minimum Wage | -0.095 | -0.041 | **-0.162*** | -0.049 | -0.041 |
|  | (-0.214 - 0.025) | (-0.339 - 0.256) | (-0.314 - -0.010) | (-0.288 - 0.191) | (-0.445 - 0.363) |
| UI | 0.028 | 0.186 | 0.061 | 0.062 | 0.215 |
|  | (-0.119 - 0.175) | (-0.052 - 0.424) | (-0.090 - 0.211) | (-0.141 - 0.266) | (-0.077 - 0.506) |
| MGS | **-0.111*** | **-0.294*** | -0.179† | -0.012 | -0.431† |
|  | (-0.220 - -0.001) | (-0.572 - -0.016) | (-0.377 - 0.019) | (-0.297 - 0.273) | (-0.862 - 0.001) |

Note: ^†^ p<0.1, ^*^ p<0.05, ^**^ p<0.01, ^***^ p<0.001

a. Panel A presents results from models where each policy variable is tested separately without controlling for state-level covariates. Panel B presents results from models similar to those in Panel A but adding state-level variables. All models are clustered at state level.

b. All = the prevalence of joint pain in states; <HS = the prevalence for the group who has less than high school degree; HS/SC= the prevalence for the group who has high school or some college degree; BA+ = the prevalence for the group who have bachelor's degree or above. SNAP = Supplemental Nutrition Assistance Program; EITC = Earned Income Tax Credit; UI = unemployment insurance; MGS = Medicaid Generosity Score.

Table S5. Coefficients of welfare policies in two-way fixed effects models with different lags of policy data.

|  | All | <HS | HS/SC | BA+ | Educational Gap |
| --- | --- | --- | --- | --- | --- |
| Panel A (no lag) |  |  |  |  |  |
| SNAP | -0.180 | -0.192 | -0.433 | -0.575 | -0.134 |
|  | (-0.551 - 0.190) | (-0.995 - 0.610) | (-1.021 - 0.155) | (-1.469 - 0.320) | (-1.171 - 0.904) |
| EITC | -0.002 | 0.001 | -0.005 | -0.031† | 0.016 |
|  | (-0.013 - 0.010) | (-0.037 - 0.040) | (-0.028 - 0.018) | (-0.064 - 0.001) | (-0.036 - 0.069) |
| Minimum Wage | -0.019 | 0.001 | -0.117 | -0.111 | 0.037 |
|  | (-0.145 - 0.107) | (-0.337 - 0.340) | (-0.308 - 0.074) | (-0.408 - 0.187) | (-0.424 - 0.497) |
| UI | 0.023 | 0.135 | 0.042 | 0.188 | 0.092 |
|  | (-0.056 - 0.102) | (-0.108 - 0.378) | (-0.125 - 0.209) | (-0.055 - 0.430) | (-0.207 - 0.391) |
| MGS | 0.026 | 0.027 | 0.093 | 0.029 | 0.021 |
|  | (-0.129 - 0.181) | (-0.327 - 0.381) | (-0.151 - 0.336) | (-0.315 - 0.372) | (-0.409 - 0.452) |
| Panel B (1-year lag) |  |  |  |  |  |
| SNAP | 0.015 | -0.432 | -0.28 | -0.389 | -0.422 |
|  | (-0.233 - 0.263) | (-1.056 - 0.193) | (-0.760 - 0.200) | (-1.150 - 0.372) | (-1.191 - 0.348) |
| EITC | 0.001 | 0 | -0.008 | **-0.021*** | 0.008 |
|  | (-0.011 - 0.013) | (-0.030 - 0.029) | (-0.021 - 0.004) | (-0.040 - -0.002) | (-0.033 - 0.049) |
| Minimum Wage | -0.061 | -0.053 | -0.137† | -0.076 | -0.058 |
|  | (-0.175 - 0.053) | (-0.352 - 0.245) | (-0.295 - 0.021) | (-0.309 - 0.156) | (-0.448 - 0.332) |
| UI | 0.047 | 0.159 | 0.081 | 0.097 | 0.176 |
|  | (-0.032 - 0.126) | (-0.041 - 0.359) | (-0.043 - 0.204) | (-0.066 - 0.261) | (-0.093 - 0.446) |
| MGS | -0.077 | -0.115 | -0.084 | 0.035 | -0.185 |
|  | (-0.203 - 0.050) | (-0.384 - 0.155) | (-0.239 - 0.071) | (-0.230 - 0.300) | (-0.550 - 0.180) |
| Panel C (3-year lag) |  |  |  |  |  |
| SNAP | -0.061 | 0.087 | -0.276 | 0.119 | 0.044 |
|  | (-0.396 - 0.273) | (-0.596 - 0.771) | (-0.613 - 0.061) | (-0.360 - 0.597) | (-0.884 - 0.972) |
| EITC | 0.008 | 0.035 | -0.011† | 0.002 | 0.042 |
|  | (-0.003 - 0.020) | (-0.009 - 0.079) | (-0.023 - 0.001) | (-0.022 - 0.026) | (-0.022 - 0.105) |
| Minimum Wage | -0.116† | -0.11 | **-0.185*** | -0.079 | -0.103 |
|  | (-0.241 - 0.008) | (-0.422 - 0.201) | (-0.348 - -0.022) | (-0.387 - 0.229) | (-0.563 - 0.357) |
| UI | 0.033 | 0.138 | 0.075 | 0.066 | 0.16 |
|  | (-0.103 - 0.168) | (-0.084 - 0.359) | (-0.071 - 0.221) | (-0.147 - 0.278) | (-0.120 - 0.440) |
| MGS | **-0.143*** | **-0.369**** | -0.210† | -0.04 | **-0.518**** |
|  | (-0.254 - -0.032) | (-0.597 - -0.142) | (-0.432 - 0.012) | (-0.389 - 0.309) | (-0.896 - -0.139) |
| Panel D (4-year lag) |  |  |  |  |  |
| SNAP | -0.032 | 0.026 | **-0.432*** | 0.016 | -0.001 |
|  | (-0.335 - 0.271) | (-0.687 - 0.739) | (-0.798 - -0.066) | (-0.547 - 0.579) | (-0.959 - 0.957) |
| EITC | 0.012† | 0.037 | -0.002 | 0.018 | 0.033 |
|  | (-0.002 - 0.027) | (-0.011 - 0.084) | (-0.025 - 0.022) | (-0.004 - 0.041) | (-0.040 - 0.106) |
| Minimum Wage | -0.073 | -0.095 | -0.071 | 0.071 | -0.191 |
|  | (-0.194 - 0.048) | (-0.352 - 0.162) | (-0.248 - 0.106) | (-0.221 - 0.363) | (-0.587 - 0.204) |
| UI | 0.025 | 0.15 | 0.021 | -0.125 | 0.219 |
|  | (-0.106 - 0.155) | (-0.060 - 0.361) | (-0.104 - 0.147) | (-0.331 - 0.081) | (-0.052 - 0.490) |
| MGS | **-0.184**** | **-0.372*** | **-0.238*** | -0.083 | **-0.503*** |
|  | (-0.308 - -0.060) | (-0.696 - -0.049) | (-0.438 - -0.038) | (-0.383 - 0.217) | (-0.965 - -0.041) |

Note: ^†^ p<0.1, ^*^ p<0.05, ^**^ p<0.01, ^***^ p<0.001

a. All models are clustered at state level and include the full set of policy variables and state-level covariates. Panels A, B, C, D represent results from models where pain outcomes are lagged by 0, 1, 3, and 4 years, respectively.

b. All = the prevalence of joint pain in states; <HS = the prevalence for the group who has less than high school degree; HS/SC= the prevalence for the group who has high school or some college degree; BA+ = the prevalence for the group who have bachelor's degree or above. SNAP = Supplemental Nutrition Assistance Program; EITC = Earned Income Tax Credit; UI = unemployment insurance; MGS = Medicaid Generosity Score.

Figure S1. Trends of prevalence changes among U.S. adults from 2011 to 2021, by state.

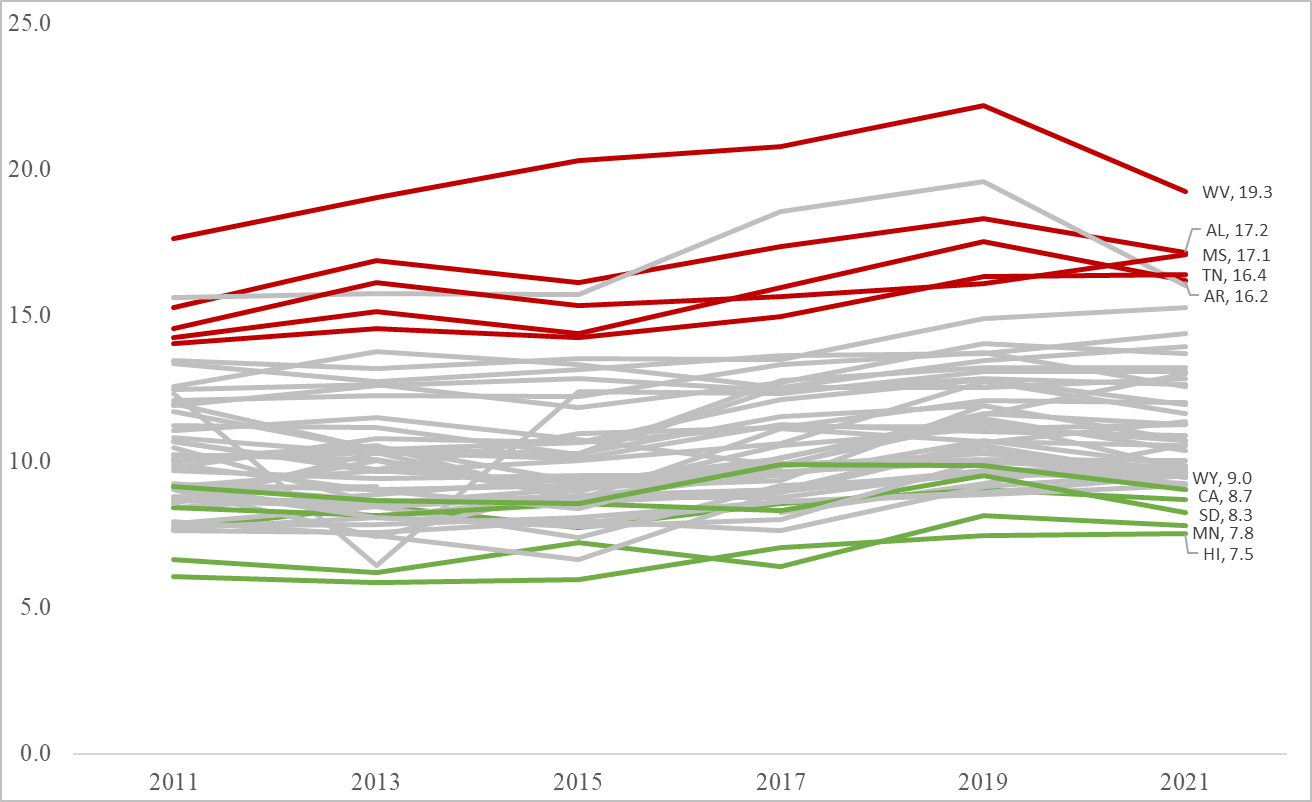

Note:

a. Data for New Jersey in 2019 and Florida in 2021 are missing as they did not meet the minimum requirements of BRFSS for inclusion.

b. Lines for states with the highest pain prevalence in 2021 are colored in red; lines for states with the lowest pain prevalence in 2021 are colored in green.

Figure S2. Trends of relative prevalence changes in educational gaps among U.S. adults from 2011 to 2021, by state.

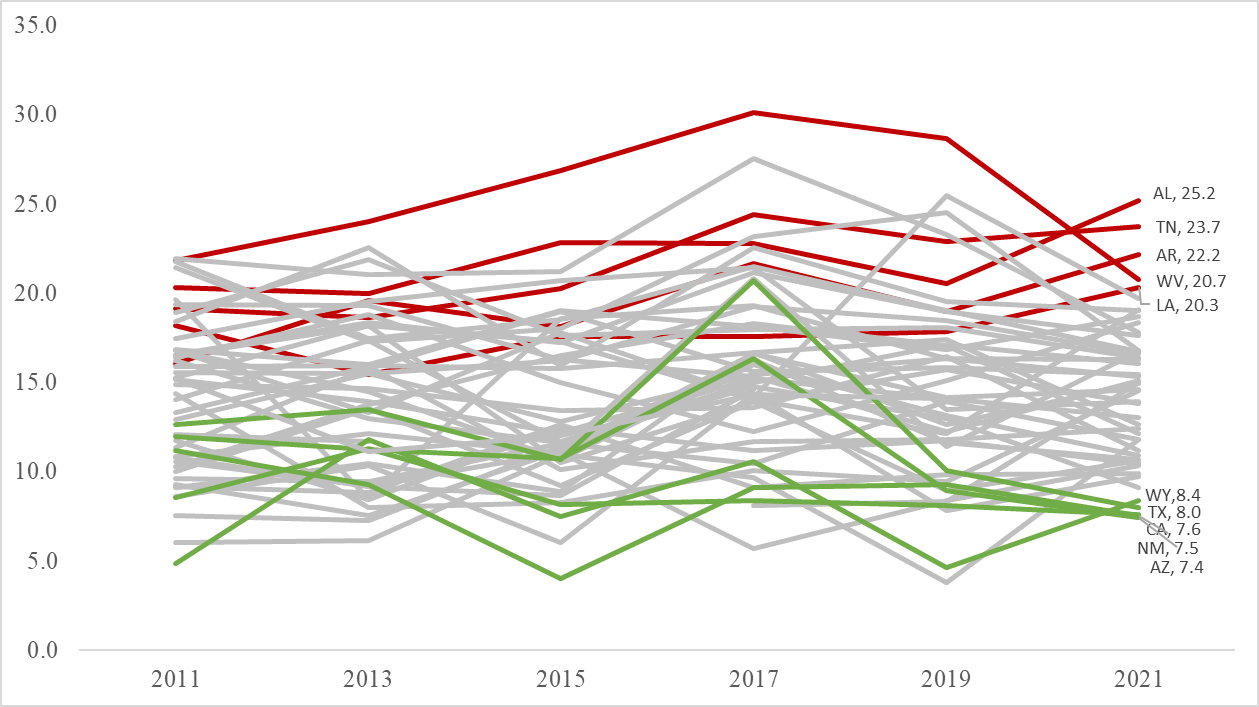

Note:

a. Data for New Jersey in 2019 and Florida in 2021 is missing as they did not meet the minimum requirements of BRFSS for inclusion.

b. Lines for states with the highest pain prevalence in 2021 are colored in red; lines for states with the lowest pain prevalence in 2021 are colored in green.
